# Ethnicity and acculturation: Asian American substance use from early adolescence to mature adulthood

**DOI:** 10.1101/19002048

**Authors:** Zobayer Ahmmad, Daniel E. Adkins

**Affiliations:** Department of Sociology, University of Utah, Salt Lake City, UT; Department of Psychiatry, University of Utah, Salt Lake City, UT

**Keywords:** Asian Americans, Population health, Substance use, Cannabis use, Social epidemiology, Risk behavior

## Abstract

Research on Asian American substance use has, to date, been limited by monolithic conceptions of Asian identity, inadequate attention to acculturative process, and a dearth of longitudinal analyses spanning developmental periods. Using five waves of the National Longitudinal Study of Adolescent to Adult Health, this study addresses these limitations by longitudinally investigating disparities in substance use from early adolescence into mature adulthood among Asian American ethnic groups, including subjects identifying as multiple Asian ethnicities and multiracial Asians. The conditional effects of acculturation indicators (e.g., nativity generation, co-ethnic peer networks, co-ethnic neighborhood concentration) on the substance use outcomes were also examined. Results indicate significant variation across Asian ethnicities, with the lowest probabilities of substance use among Chinese and Vietnamese Americans, and the highest among multiracial Asian Americans. Acculturation indicators were also strongly, independently associated with increased substance use, and attenuated many of the observed ethnic disparities, particularly for multiracial, multiethnic, and Japanese Asian Americans. This study argues that ignoring the diversity of Asian ethnicities masks the presence of high-risk Asian American groups. Moreover, results indicate that, among Asian Americans, substance use is strongly positively associated with acculturation to U.S. cultural norms, and generally peaks at later ages than the U.S. average.

---

Recent research into the social epidemiology of substance use has generated important insights into the impact of substance misuse on U.S. life expectancy, and social disparities in morbidity and mortality within the U.S. population (Case and Deaton 2015, Ruhm 2017, Ruhm 2018). Such research has advanced understanding of the relationship of substance use to morbidity and mortality patterns, particularly among non-Hispanic White Americans (Geronimus et al. 2019, Shanahan et al. 2019). However, knowledge regarding the social epidemiology of substance use among other racial/ethnic groups in the U.S., particularly Asian Americans (AAs), remains limited. This is in spite of notable recent growth and increasing diversity in the AA population (Cook et al. 2015, Saraiya et al. 2019).

While it is established that AAs exhibit a relatively healthy substance use profile in aggregate (Martell, Garrett and Caraballo 2016), studies disaggregating AA ethnicities have also suggested the presence of high risk Asian subgroups (Cook et al. 2015, Saraiya et al. 2019, Thai, Connell and Tebes 2010). For instance, Korean and Japanese ethnicities appear to have substantially higher rates of alcohol use than other AA ethnicities (Iwamoto, Takamatsu and Castellanos 2012, Tosh and Simmons 2007). Moreover, the prevalence of cigarette smoking among Koreans in the U.S. was 20% in 2013, a rate comparable to other high-risk U.S. demographic groups (Martell, Garrett and Caraballo 2016).

Extant research on substance use among AAs has primarily focused on tobacco and alcohol use, with few studies examining illicit substance use (e.g., cannabis, opioids, and illicit stimulants) (Cochran et al. 2007, Hong et al. 2011, Liu and Iwamoto 2007, Saraiya et al. 2019, Yoo et al. 2010). The small body of research examining illicit substance use across AA ethnicities has indicated the presence of significant ethnic variation. For instance, cross-sectional studies have shown AA ethnic variation in illicit substance use rates, with Filipino, Japanese, and Korean ethnicities reporting more use than their Asian peers (Ryabov 2015, Saraiya et al. 2019). Similarly, using data from Waves 1 and 3 of the National Longitudinal Study of Adolescent to Adult Health, Ryabov (2015) found that substance use rates, especially the use of illicit drugs, vary widely across Asian ethnicities in adolescence and early adulthood (e.g., youth of Filipino ethnicity exhibit higher prevalence of illicit drugs than East Asian peers (Ryabov 2015)).

Despite the notable demographic growth of AAs identifying as multiethnic (e.g., Vietnamese and Chinese) and multiracial (e.g., Vietnamese and White), to date, very little research has examined substance use among these increasingly common ethnic identities. Moreover, existing studies offer minimal sociocultural explanation for substance use disparity across Asian ethnicities (Dunbar et al. 2018, Price et al. 2002, Wong, Klingle, and Price 2004). To address these shortcomings, we analyze five waves of the National Longitudinal Study of Adolescent to Adult Health (Add Health) to assess patterns of substance use disparities among AAs, and determine to what extent these disparities are explained by variation in acculturation to American norms. Further, we also map developmental patterns in AA substance use, going beyond cross-sectional data (and longitudinal data limited to a single developmental period) to examine longitudinal AA ethnic variation in substance use from early adolescence (age ~12) to mature adulthood (age ~40). The wide age range of Add Health allows detailed description of longitudinal AA substance use patterns, revealing persistent disparities across AA ethnicities and nativity generations, and unique developmental patterns among AAs relative to other U.S. racial/ethnic groups.

## Background

### Diverse sociodemographic profiles of Asian Americans

The Asian population is the fastest growing of all racial/ethnic groups in the U.S., driven by consistently high immigration and relatively high fertility rates (López, Ruiz and Patten 2017). The size of the Asian population more than doubled (from 11.9 million to 20.4 million) between 2000 and 2015 (López, Ruiz and Patten 2017). Among the 19 national origins that comprise 94 percent of U.S. Asian population, the largest groups are Indian, Chinese, and Filipino origin. Along with demographic and underlying sociocultural heterogeneity, there is also marked socioeconomic inequality within the AA population, despite the general perception of Asians in the U.S. as high achieving (Lee 2015). Monolithic conceptions of AA identity, combined with the model minority stereotype, has served to mask socioeconomic inequality across AA individuals and ethnicities. For instance, recent estimates suggest that Asians who are in the top 10% of the income distribution earn >10x more than Asians who are in the bottom decile, making Asians the most economically unequal racial/ethnic group in the U.S. (Kochhar and Cilluffo 2018). Moreover, there are substantial ethnic difference across AA income deciles, with South and East Asian ethnicities exhibiting socioeconomic advantage relative to other AA ethnicities (Cook et al. 2017). Given the continuing demographic, cultural, and economic diversification of the AA population, scholars have called for increased attention to ethnic-specific health and health behavior patterns (Yoo, Le and Oda 2013).

Demographic literature indicates that marriage between two Asian ethnicities, as well between Asians and other racial/ethnic groups is rising (Lee and Bean 2004, Qian and Lichter 2011). As a result, there is an increasing number of individuals identifying as multiple Asian ethnicities (e.g., Vietnamese and Chinese), and individuals identifying as multiracial (e.g., Japanese and White). Despite this growth in multiethnic and multiracial Asian populations, there is virtually no research investigating the patterns and prevalence of substance use among these groups, as they are frequently classified as mono-ethnic and/or mono-racial. This failure to consider multiethnic and multiracial Asian identities may confound inferences regarding the relationships of ethnicity and substance use. Thus, we give explicit consideration to multiethnic and multiracial AAs, along with the surveyed mono-ethnic Asian identities.

### Variation in substance use patterns in nations of origin

Despite widespread perceptions to the contrary, not all Asian ethnic groups benefit equally from salubrious heritage cultural norms against substance use. Some types of substance use, particularly alcohol use and tobacco smoking, remain significant public health challenges in Asian nations of origin (Cook, Mulia and Karriker-Jaffe 2012, Cook et al. 2013, Cook et al. 2015). Cook et al. (2013) show that it is not only a loss of heritage culture, but also diverse Asian ethnic drinking cultures in their nations of origin that are associated with elevated alcohol use patterns in the U.S.. According to recent reports,

Korea and Japan have the highest alcohol consumption prevalence in Asia, as measured by WHO per capita annual alcohol consumption (Cook et al. 2013). Moreover, many Asian heritage cultures are characterized by highly gendered patterns of substance use, characterized by much higher substance use among males relative compared to females (Li and Wen 2015). This pattern is particularly evident in many East and Southeast Asian nations, such as South Korea, Philippines, and China, where tobacco smoking prevalence rates are ≥40% for males and <10% for females (WHO 2019).

### Acculturation and substance use

Acculturation refers to post-immigration changes in immigrant individuals’ way of life, including changes in physical, psychological, and cultural traits due to continuous exchange with the new sociocultural environment (Le, Goebert and Wallen 2009). Behavioral changes that take place during acculturation remain a primary focus in immigrant health literature (e.g., Abraído-Lanza, Chao and Florez 2005). Sociologists have theorized that behavioral changes, such as the adoption of regular alcohol consumption or transitioning to fast food culture, are largely influenced by the length of U.S. residence—that is, the longer the duration of residence in the U.S., the greater the extent of behavioral changes (Yoo, Le and Oda 2013). Previous research indicates that English language use, particularly in personal conversation, is associated with behavioral risk-taking, such as heavy drinking and misuse of prescription drugs (Bersamira et al. 2017, Park et al. 2014).

Research on AA substance use, mainly alcohol consumption and tobacco use, has commonly examined the effects of acculturation—which is typically measured by nativity, language use, duration of stay in the U.S. Previous research has shown that acculturation predicts increased substance use (Becerra et al. 2013, Iwamoto et al. 2016, Le, Goebert and Wallen 2009, Lui and Zamboanga 2018), suggesting acculturation as a risk factor for AA health and health behavior (Le, Goebert and Wallen 2009). For instance, research using the National Latino and Asian American Study (NLAAS) found that nativity generation in the U.S., and English language use were positively associated with alcohol consumption, cannabis, cocaine, and other drug use (Bersamira et al. 2017). A systematic literature review of 31 studies on the relation between acculturation and alcohol consumption found that acculturation is a primary mediator of hazardous drinking behavior among AAs (Lui and Zamboanga 2018). Less is known of how contextual acculturation factors, such co-ethnic friendship networks and neighborhood composition may influence substance use. Further, it is unclear what role acculturation may play among multiethnic and multiracial Asians, who are may be subject to uniquely racialized acculturative processes (Schwartz et al. 2010).

Empirical research examining the effects of acculturation on substance use among AA adolescents born into immigrant families is lacking. However, family is clearly a primary influence promoting the retention of heritage culture among first and second generation AAs. It is documented that Asian-born parents actively socialize their children in heritage values and culture, with special emphasis on adolescent behavioral conformity to rules and norms (Nelson et al. 2015). This suggests highly acculturated AAs (3+ generation, multiracial) may be at greater behavioral risk, as they lack the buffering socialization associated with first or second generation status. Support for such acculturation effects come from a recent, large meta-analysis (N = 68,282) demonstrating that attachment to Asian heritage culture is negatively associated with alcohol consumption (Lui and Zamboanga 2018). Thus, we hypothesize parental nativity, as well as respondents’, are especially important distinctions for ethnic groups comprised primarily of recent immigrants (e.g., Chinese, Vietnamese) (Le and Kato 2006, Telzer, Gonzales and Fuligni 2014).

It has also been noted that peer homophily among AA immigrants may buffer against risk behavior. For instance, analysis of early waves of Add Health indicates that having close friends from similar ethnic background and nativity status is protective against illicit drug use among AAs, suggesting a salubrious influence for immigrant and co-ethnic social network ties (Ryabov 2015). To assess the role of acculturation in personal relationships, we include measures of co-ethnic peer friendship network composition, co-ethnic neighborhood density, and English use in familial/private conversations in this analysis.

### Developmental variation in substance use

Another limitation of in the social epidemiology of substance use literature is overreliance on cross-sectional data and adolescent samples. This is a notable limitation, as it is well established that substance use and misuse is not evenly distributed across age. Previous research has suggested that, in contemporary U.S. cohorts, substance use patterns are characterized by a sharp rise in late adolescence and young adulthood, followed by a decline in mature adulthood (i.e., 30s) (Chen and Jacobson 2012, Fothergill et al. 2016, Merline et al. 2004). Analysis of longitudinal data is therefore essential, particularly so among AAs, as previous research has also shown that AAs may show atypical patterns of externalizing, and internalizing, traits during adolescence and the transition to adulthood, likely due to particularly restrictive family environments in adolescence, before converging toward mainstream U.S. cultural norms in young adulthood ([Author(s)], Hahm et al. 2014). Findings from a study on substance use among Asian and Pacific Islander sexual minorities also support this pattern, showing that the association between sexual minority status and substance use does not emerge until young adulthood (Hahm et al. 2008), suggesting young adulthood as a period of fuller expression of AA behavioral preferences. Thus, exiting from parental supervision during adulthood may lead unique developmental patterns of substance use among AAs, likely characterized by substance use peaking at later ages than the U.S. average. To address this gap in the literature, we examine developmental trends in substance use across AA ethnicities.

### Research questions

This study investigates substance use variations across AA ethnicities, including multiethnic and multiracial AAs, and examines potential mediation of ethnic disparities by acculturation status. Additionally, it examines ethnic variation in developmental patterns of substance use from adolescence to mid-adulthood (ages ~15-40) using Waves 1-5 of the National Longitudinal Study of Adolescent to Adult Health. Specific hypotheses include:

*Hypothesis 1:* There will be significant ethnic heterogeneity in substance use rates. AA groups with weaker attachment to traditional Asian norms due to (a) diverse family composition (i.e., multiethnic and multiracial Asians), and (b) longstanding residence in the U.S. (i.e., Japanese) will be more likely to use all four substance use outcomes (tobacco, alcohol, cannabis, and illicit substances). *Hypothesis 2:* The relationship between ethnicity and substance use will be substantially attenuated by conditioning on acculturation indicators, including generational status, English language use in familial/private settings, and co-ethnic peer network and neighborhood composition. *Hypothesis 3:* The prevalence of substance use in this longitudinal cohort of AAs will peak after the normative peak, age ~25, in contemporary U.S. society.

## Materials and Methods

### Data

Data for the project come from the National Longitudinal Study of Adolescent to Adult Health (Add Health) Wave 1-Wave 5. Add Health is a large, nationally representative, longitudinal study that began in 1994, now including five waves of data spanning 28 years of data collection. Add Health collected detailed information on environmental, behavioral, and biological information (Entzel and Udry 2009). Wave 1 of Add Health study surveyed about 90,000 adolescents across 80 high schools and 52 middle schools throughout the United States. Out of the total sample, a subsample of 20,747 students and their parents filled out an in-depth home interview survey. Wave 2 of Add Health was completed in 1996, containing only the students who were in 8^th^-12^th^ grades (excluding senior students) (N=14,738). A sample of 15,197 young adults and their partners (ages 18-26) responded in Wave 3 survey. Wave 4 collected data from 15,701 adults (ages 24-32), and Wave 5 surveyed 12,300 subjects (ages 32-42). As the current study is focused on within Asian subgroup differences, we only analyzed AAs from Add Health. These included 1585 AAs in Wave 1, following up 1089, 1195, 1060, and 857 in Waves 2, 3, 4, and 5, respectively.

### Measures

Due to cultural differences in Asian nations of origin, and segmented assimilation in the U.S., there is reason to suspect variable ethnic disparities across substances (e.g., relatively high rates of smoking among Koreans, and drinking among Japanese) (Iwamoto et al. 2016, Tosh and Simmons 2007, Yoo et al. 2010). Thus, we adopt a comprehensive approach to modeling substance use, examining 4 outcomes: tobacco, alcohol, cannabis, and illicit substances.

#### Substance use

Dependent variables come from Wave 1 to Wave 5 of Add Health. Information on tobacco smoking, alcohol consumption, and cannabis use were asked in each of the 5 waves of data collection. Specifically the items used here are: 1) *Ever regular tobacco smoker*: has the subject ever smoked cigarettes regularly (at least 1 cigarette every day for 30 days); 2) *Sustained regular alcohol use:* has the subject regularly consumed alcohol in the past year (≥1-2 days per week); 3) *Recent (past month) cannabis use:* has the subject consumed cannabis in the past month.; 4) *Recent (past month) illicit substance use*: how many of the following substance use categories has the subject used in the past month, a) cocaine/crack; b) inhalants/methamphetamine, and c) other illicit drugs (e.g., illicit prescription drug use [e.g., opioid, stimulant, benzodiazepine], heroin, and psychedelic drugs (e.g., LSD, psilocybin, PCP)). The decision to pool the illicit substances into a count variable was guided by the very low base rates for the individual drugs, and also by psychometric evidence from previous research of a general propensity to illicit drug use (Connor et al. 2014).

#### Ethnicity

A primary objective of the current study is to examine substance use variation across Asian ethnic groups, to unveil the heterogeneity masked by the model minority stereotype. Fortunately, Add Health sampled key Asian ethnicities including Japanese, Filipino, Chinese, Vietnamese, Indian, and Korean. Information on racial/ethnic identity was derived from a set of questions querying whether the subject identifies as Asian and, if so, which Asian ethnicities. Response options to the item included Japanese, Filipino, Chinese, Vietnamese, Asian Indian, and Korean, as well as “Other” (i.e., Asian, ethnicity not specifically reported). The “Other” Asian category includes respondents identifying any Asian country other than the six countries explicitly specified, likely yielding a heterogeneous group consisting substantially of individuals of Pakistani and Southeast Asian (e.g., Cambodian, Hmong, Thai) descent. Additionally, some respondents identified as more than one Asian ethnicity (e.g., Japanese and Korean), but no other races, which are categorized as “multiethnic Asians.” Finally, some individuals of Asian descent also identified as multiple racial groups (e.g., Asian and White), and are categorized as “multiracial Asian”. Ethnic categories are coded mutually exclusively.

#### Sociodemographics

Models also include a set of household SES measures for parental education and household income assessed at Wave 1. We used the parental questionnaire to ascertain the educational attainment of the sampled residential parent (typically the mother) and current partner (typically the father). This scale ranged 0-9, with 0 corresponding to no schooling, and 9 corresponding to “professional training beyond a four-year college or university”. Parental educational attainments were averaged across parents, when more than one was reported. Household income was ascertained from the parental questionnaire and included all sources of income from the previous year (measured in thousands of dollars) and is natural log transformed to reduce pronounced skew [Author(s)].

#### Acculturation

Measures of acculturation include respondents’ nativity status, parental nativity status, English language use in private/familial settings, co-ethnic friend network proportion, and co-ethnic neighborhood (census tract) proportion. Information on the subject’s nativity status, and their parents’ nativity status come from Wave 1. Using three items (subject, mother, and father’s nations of origin), we classified subjects’ generational status as 1, 2, or 3+. *Generation 1* includes foreign-born respondents with both parents also foreign-born; *generation 2* includes U.S.-born children with two foreign-born parents; *generation 3+* consists of U.S.-born children with U.S.-born parent/s. English language use in familial/private settings was assessed in Waves 1-3. This measure was estimated in Waves 4 and 5 as the average of the preceding waves. Co-ethnic friend network proportion was estimated from the Wave 1 in-school friend nomination data. In the in-school friendship data, subjects nominated up to 10 friends, which were matched with students of known race in the in-school sample, thus allowing estimation of co-ethnic friendship network proportion (= # known-Asian friends / total # known-race friends). Finally, co-ethnic neighborhood (census tract) proportion was assessed by matching subjects to tract-level census/American Community Survey (ACS) data in Waves 1-4, with Wave 5 estimated as the average of the preceding waves.

### Statistical Analysis

#### Descriptive statistics and visualization

The intersecting effects of ethnicity and acculturation on substance use patterns is a primary interest of this project. We visualize these relationships in Figure 1 using a series of bar graphs visualizing variation in substance use across nativity generation, within each ethnicity. Specifically, bar graphs showing mean tobacco, alcohol, cannabis, and illicit substance use for each ethnicity-generation combination, with 95% confidence interval bars. For each substance, ethnicities are ordered in ascending mean substance use prevalence; 95% CIs were calculated using cluster (observations within subjects) robust SEs (MI=30). Additionally, as substance use patterns are highly gendered (e.g., McHugh et al. 2018), we also present bar graphs showing mean tobacco, alcohol, cannabis, and illicit substance use across Asian ethnicities and sex, with 95% confidence interval bars in Figure 2.

Given the well-documented patterns in substance use rates across developmental periods (e.g., elevated rates during late adolescence and young adulthood), we include a visualization of AA ethnic differences in mean substance use across the age range (ages ~15-40) in Figure 3. We restrict this investigation to a descriptive visualization of ethnic variation in age patterns of substance use, as the sample is not adequately powered to conduct inferential tests of higher order interactions (e.g., age^2^×ethnicity) for the smaller ethnic subsamples (e.g., Asian Indian, Vietnamese). Thus, for each of the four substance use outcomes, we visualize smoothed mean levels of substance use over age, disaggregated by AA ethnicities, using median spline plots (i.e., cubic regression splines with cross-median knots), specifying four knots to smooth age-by-ethnicity bins with small cell sizes (Royston and Sauerbrei 2007). Additionally, in Figure 4 we visualize smoothed mean levels of the four substance use outcomes over age, disaggregated into the six nativity generation-sex combinations, using the median spline method described above.

#### Inferential analyses

Primary inferential analyses examine four outcomes using generalized linear mixed models to jointly analyze the Asian subsample across Waves 1-5 of Add Health (N=7920). Three of the four outcomes are dichotomous: (a) ever daily tobacco smoking, (b) sustained regular alcohol consumption (≥1-2 per week, previous year), and (c) smoking cannabis in the past month. Thus, these 3 outcomes use a logistic link function. The last outcome, (d) illicit substance use in the past month, is a count variable of the number of illicit substance categories used in the past month (0-3); it was analyzed with a negative binomial link function given over-dispersion. All models specified a subject-level random intercept to account for the multilevel data structure in which observations are nested within subjects. Among other desirable properties, this approach corrects standard errors and coefficients for nonindependence of observations within subjects (Gelman and Hill 2006).

Each of the four outcomes followed the same sequence of four model specifications. In Model 1 of the sequence, the outcome was predicted with only the ethnicity indicators, which categorize Asian ethnicities into eight mutually exclusive groups. Thus, the first model assesses unconditional differences in substance use across Asian ethnic groups. Model 2 of the sequence includes the ethnic indicators from Model 1 and adds controls for other sociodemographic variables—age, age^2^, sex, parental education, and childhood household income. Model 3 included all variables from Model 2, as well as contextual acculturation factors (i.e., co-ethnic neighborhood composition, and co-ethnic friendship network proportion). Models 4 includes all measures from Model 3, as well as generational acculturation factors (i.e., nativity generation 1 (ref), 2, and 3+, and use of English in private/familial settings). Results are reported as coefficients (i.e., logged odds ratios in the logistic models, and logged incidence rate ratio in the negative binomial model) with t statistics in parenthesis. Given that our analysis is limited to Asian ethnicities only, the analysis is, by design, not nationally representative. Considering this along with the observation that, due to the oversampled nature of Asian subsample in Add Health, using person weights may diminish inferential efficiency (Bollen et al. 2016), our primary analyses are unweighted. Sensitivity analysis adjusting for school-based PSUs and region strata did not affect substantive results.

Finally, based on the age and age squared estimates from the final models of the series, we estimated the average age of peak use for each of the four substance use outcomes as a nonlinear combination of parameter estimates, using the quadratic vertex formula: *v* = - *β_age_* / (2×*β_age_*_^2_). Standard errors for the age peaks point estimates were calculated using the delta method (Oehlert 1992), by the Stata post-estimation command *nlcom*. All analyses performed using Stata 15.1.

#### Missing data

Missing data is an ubiquitous issue longitudinal survey data (Kline, Andridge and Kaizar 2017), and is present to a modest degree in the current analysis (see Appendix 1 for missingness summary).

Conventional listwise deletion (i.e., complete case analysis) methods for addressing missing data are known to, on average, increase bias in coefficients under missing at random (MAR) and missing completely at random assumptions (MCAR), and to weaken statistical power, generally (White, Royston and Wood 2011). To avoid these potential biases and maximize statistical power, we handle missingness using *multiple imputations by chained equations* (MICE) method for the full Add Health sample (N=20,774). MICE is a statistical approach to impute values for the missing data based on distributions of the observed data, using MCMC iterative methods. This is a well-established, validated approach to multiple imputation, which preserves the distributions (i.e., continuous, count, ordinal, categorical) of the imputed variables. The performance of MICE has been demonstrated equivalent to the conventional multivariate normal multiple imputation method (Lee and Carlin 2010). All multiple imputation analyses included a conservative 30 imputations (MI=30) (von Hippel 2018).

## Results

### Descriptive analyses

Table 1 shows the descriptive statistics of the full analysis sample. The proportion of respondents reporting ever daily smoking tobacco, regularly drinking alcohol, and using cannabis in the past month, averaged across waves, are 26%, 18%, and 12%, respectively. The mean number of illicit substances used in the past month was 0.09. Among mono-ethnic Asian ethnicities, Filipino ethnicity comprise 34% of the sample, followed by Chinese at 17%. Multiracial and multiethnic Asians comprise 17% and 5% of the sample, respectively. The average age of the respondents is 25.3, with a range of 12 to 43 years. Almost half of the respondents are women (48%). The percent of the sample identifying as generation 1 (foreign-born) is 37%, generation 2 (U.S.-born, parents foreign-born) is 28%, and generation 3+ (U.S.-born, parent/s U.S.-born) is 35%. Two thirds of observations (66%) reported regular use of English in familial/private conversation.

**Table 1:** Descriptive statistics of the analysis sample. Standard deviations (SD) given for non-dichotomous measures and number of subject endorsing category (n) given for dichotomous measures. (MI=30)

| Variable | Mean | SD/n | Min | Max | N |
| --- | --- | --- | --- | --- | --- |
| **<u>Substance use</u>** |  |  |  |  |  |
| Tobacco (ever daily smoker) | 0.26 | 2059 | 0 | 1 | 7920 |
| Alcohol (regular, past year) | 0.18 | 1426 | 0 | 1 | 7920 |
| Cannabis (any in past month) | 0.12 | 950 | 0 | 1 | 7920 |
| Illicit subs. (count, past month) | 0.09 | 0.34 | 0 | 3 | 7920 |
| **<u>Ethnicity</u>** |  |  |  |  |  |
| Chinese | 0.17 | 1340 | 0 | 1 | 7920 |
| Filipino | 0.34 | 2725 | 0 | 1 | 7920 |
| Japanese | 0.03 | 200 | 0 | 1 | 7920 |
| Asian Indian | 0.02 | 155 | 0 | 1 | 7920 |
| Korean | 0.06 | 450 | 0 | 1 | 7920 |
| Vietnamese | 0.05 | 385 | 0 | 1 | 7920 |
| Other Asian | 0.11 | 885 | 0 | 1 | 7920 |
| Multietnic Asian | 0.05 | 430 | 0 | 1 | 7920 |
| Multiracial Asian | 0.17 | 1350 | 0 | 1 | 7920 |
| **<u>Sociodemographics</u>** |  |  |  |  |  |
| Age | 25.3 | 8.3 | 12 | 43 | 7920 |
| Sex (Female) | 0.48 | 3802 | 0 | 1 | 7920 |
| Parental Education (Mean) | 6.23 | 2.13 | -0.31 | 11.14 | 7920 |
| Household Income (Logged 1000s) | 3.67 | 0.84 | 0 | 6.79 | 7920 |
| **<u>Acculturation Indicators</u>** |  |  |  |  |  |
| Generation 1 | 0.37 | 2925 | 0 | 1 | 7920 |
| Generation 2 | 0.28 | 2230 | 0 | 1 | 7920 |
| Generation 3 | 0.35 | 2765 | 0 | 1 | 7920 |
| English Use | 0.66 | 5227 | 0 | 1 | 7920 |
| Co-ethnic tract proportion | 0.25 | 0.21 | -0.14 | 0.89 | 7920 |
| Co-ethnic friend network | 0.62 | 0.35 | -0.07 | 1.46 | 7920 |

Table 2 presents a bivariate frequency table of generational status by Asian ethnicity. Among Asian ethnicities, Vietnamese have the highest percentage of first generation (70.1%), followed by Korean (54.4%), Filipino (50.6%), and Chinese (41.4%). Notable, the vast majority of Japanese subjects are 3+ generation (92.5%), and no Japanese Americans in the sample identify as first generation. A large majority of multiethnic Asians (70.9%) and multiracial Asians (81.9%) are also 3+ generation. Almost half (45.2%) of Asian Indians identify as second generation, and notably few Asian Indians identify as 3+ generation (12.9%).

**Table 2:** Distribution of generation status by Asian ethnicities, with row percentages. (MI=30)

| **Ethnicity** | **<u>Generation 1</u>** |  | **<u>Generation 2</u>** |  | **<u>Generation 3</u>** |  | **N** | **Obs** |
| --- | --- | --- | --- | --- | --- | --- | --- | --- |
|  | **N** | **%** | **N** | **%** | **N** | **%** |  |  |
| Chinese | 111 | 41.4 | 106 | 39.6 | 51 | 19 | 268 | 1340 |
| Filipino | 276 | 50.6 | 190 | 34.9 | 79 | 14.5 | 545 | 2725 |
| Japanese | 0 | 0 | 3 | 7.5 | 37 | 92.5 | 40 | 200 |
| Asian Indian | 13 | 41.9 | 14 | 45.2 | 4 | 12.9 | 31 | 155 |
| Korean | 49 | 54.4 | 11 | 12.2 | 30 | 33.3 | 90 | 450 |
| Vietnamese | 54 | 70.1 | 17 | 22.1 | 6 | 7.8 | 77 | 385 |
| Other Asian | 47 | 26.6 | 66 | 37.3 | 64 | 36.2 | 177 | 885 |
| Multiethnic | 13 | 15.1 | 12 | 14 | 61 | 70.9 | 86 | 430 |
| Multiracial | 22 | 8.1 | 27 | 10 | 221 | 81.9 | 270 | 1350 |
| Total | 585 | 100 | 446 | 100 | 553 | 100 | 1,584 | 7920 |

Figure 1 visualizes mean tobacco, alcohol, cannabis use, and illicit substance use across Asian ethnicities and nativity generations, with 95% CI bars. The ethnicities are ordered in ascending mean substance use prevalence. There are notable substance use variations across Asian ethnicities, some consistent across outcomes, and some variable. In general, multiracial Asians, Asian Indians, and multiethnic Asians are show a pattern of higher rates of substance use, and Chinese and Vietnamese show the lowest rates substance use, among the Asian ethnicities. The figure supports expectations of relatively elevated smoking (Korean) and drinking (Japanese) ethnic pattern. Asian Indians exhibit an interesting pattern characterized by very low tobacco smoking rates, but relatively high alcohol, illicit, and cannabis use estimates. It is important, however, to note the relatively large confidence intervals for Asian Indians and, thus, the imprecision of these estimates. There is also a clear visual trend in Figure 1 of increasing substance use across nativity generations (with the notable exception of illicit substances), with more native generations generally exhibiting higher substance use rates, consistent with inferential results in Tables 3 and 4.

**Figure 1:**
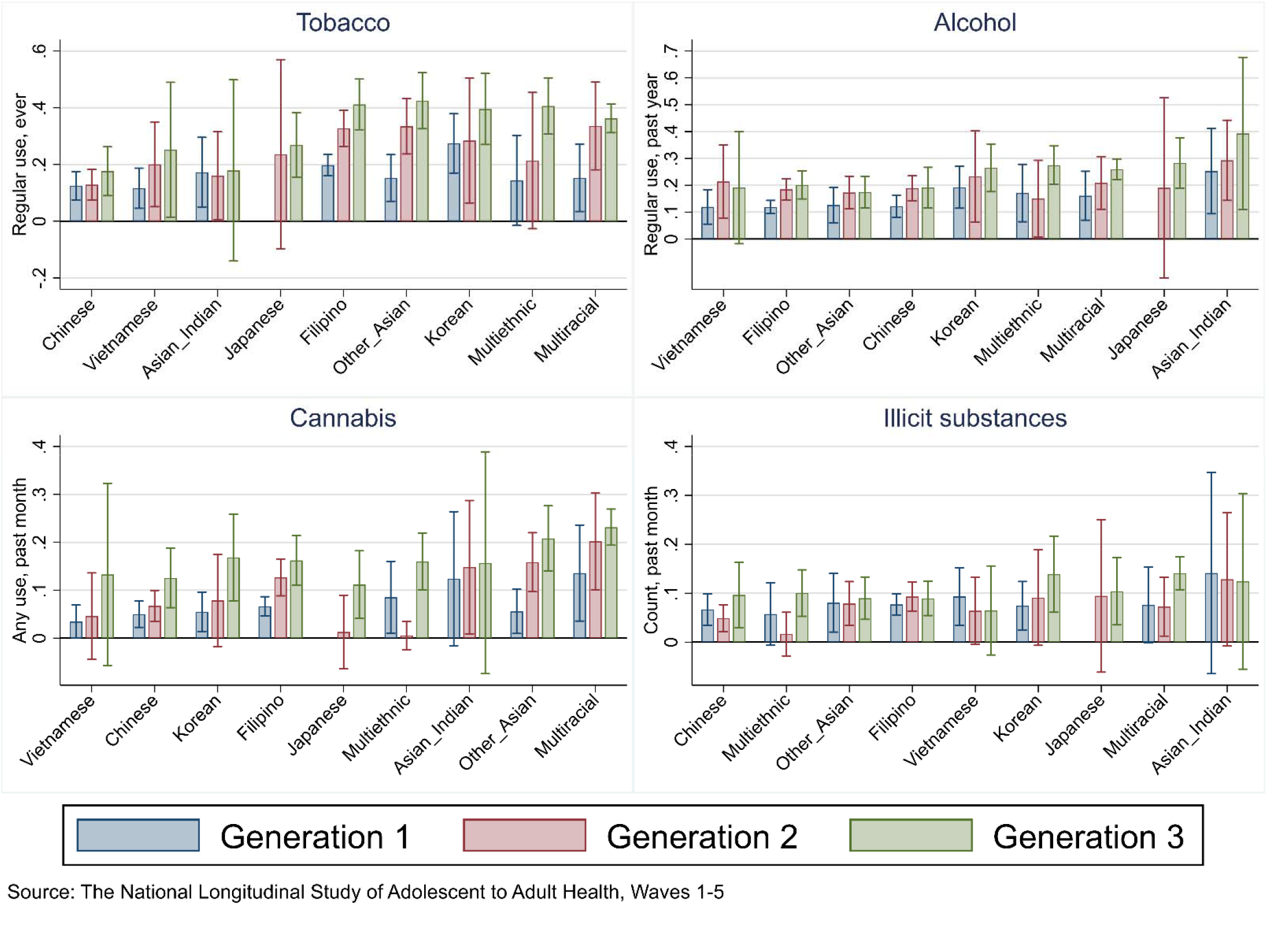
Bar graphs showing mean tobacco, alcohol, cannabis, and illicit substance use across Asian ethnicities and immigrant generations, with 95% confidence interval bars. For each substance, ethnicities are ordered in ascending mean substance use prevalence. CIs were calculated using cluster (obs within subjects) robust SEs (MI=30)

Figure 2 visualizes mean tobacco, alcohol, cannabis use, and illicit substance use across Asian ethnicities and sex, with 95% CI bars. The ethnicities are ordered in ascending substance use prevalence. The pronounced sex disparity in substance use is evident in all panels of Figure 2, with males exhibiting substantially increased levels of substance use across substances and ethnicities. Filipino Americans consistently have relatively large sex gaps in substance use, while Chinese American exhibit little sex disparities (and uniformly low levels across substances and sex). Additionally, there are more granular disparities indicated in Figure 2, including large sex disparities among Asian Indian and Vietnamese ethnicities in tobacco use, and among Japanese ethnicity in alcohol and cannabis use.

**Figure 2:**
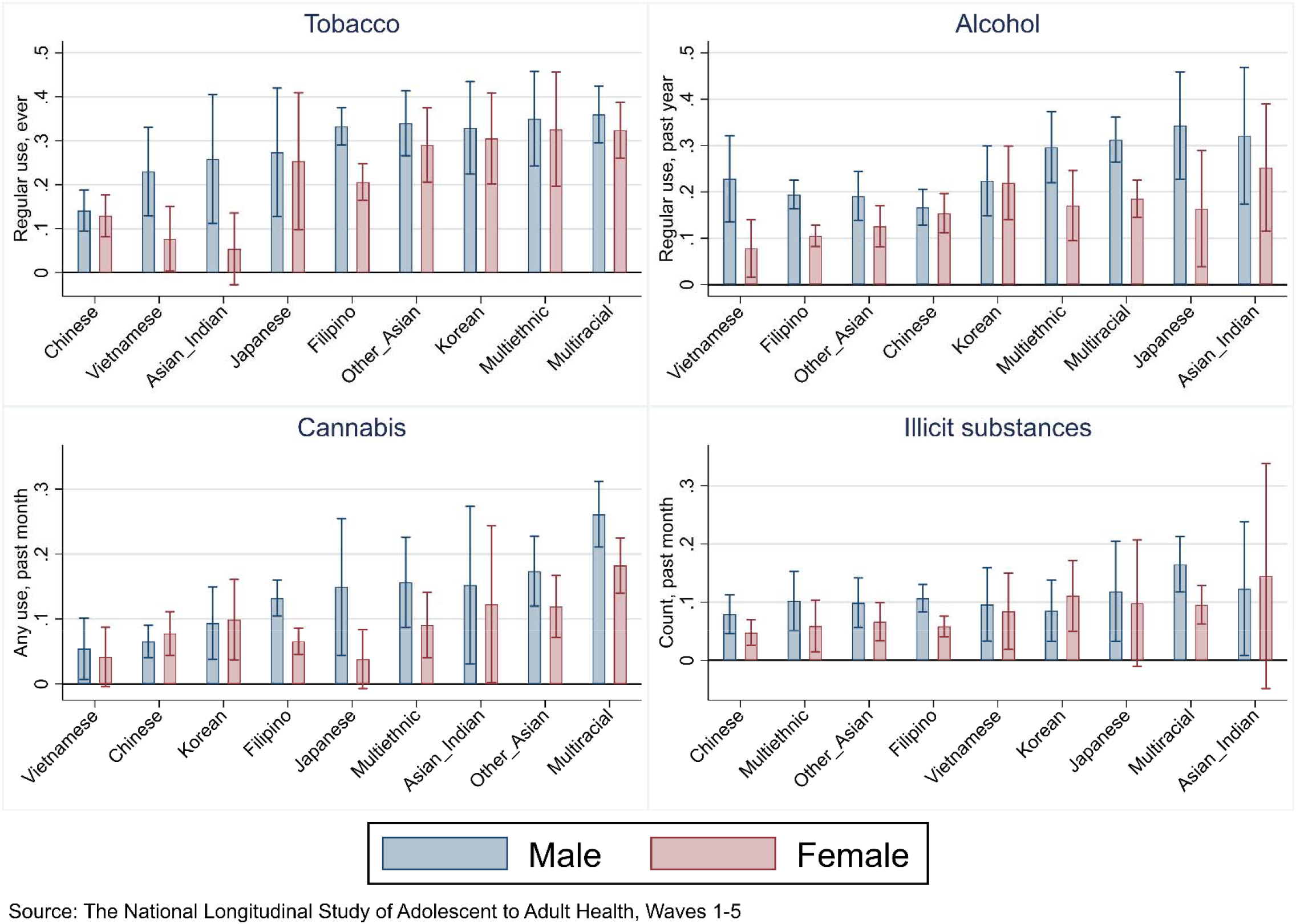
Bar graphs showing mean tobacco, alcohol, cannabis, and illicit substance use across Asian ethnicities and sex, with 95% confidence interval bars. For each substance, ethnicities are ordered in ascending mean substance use prevalence. CIs were calculated using cluster (obs within subjects) robust SEs (MI=30)

Figure 3 visualizes smoothed substance use means by age and Asian ethnicity for tobacco, alcohol, cannabis, and illicit substances. Due to some small age-ethnicity cells, we use median splines (with four knots) to smooth means over age (Wegman and Wright 1983). Asian ethnicities vary in terms of growth and decline in rates of substance use as the subjects age from ~15-40. We observe some general age trends that are fairly consistent across ethnicities. For instance, the age pattern of ever regular tobacco use indicates a marked increase in the teens and early/mid-20s before stabilizing in the late 20s. Conversely, current regular alcohol tends to stabilize in either the mid-20s (e.g., multiracial, multiethnic, and Japanese) or the early 30s (e.g., Asian Indian, Chinese, Vietnamese). Cannabis use in the past month is characterized by heterogeneity across Asian ethnicities, with most ethnicities peaking in the early/mid-20s, declining in the early/mid-30s, and then rising again in the late 30s. An exception to this pattern is multiracial Asians, who exhibit a sustained relatively very high (though declining with age) level of cannabis use throughout the age range. Finally, illicit substance use show uniformly low levels across ethnicity, but also a notable pattern of growth over the longitudinal age range, steadily increasing from the mid-teens to the early 40s. Ethnic differences mirror the results in Figures 1 and 2, with Chinese and multiethnic Asians typically at the lowest levels across the age range, and multiracial Asians at the highest levels.

**Figure 3:**
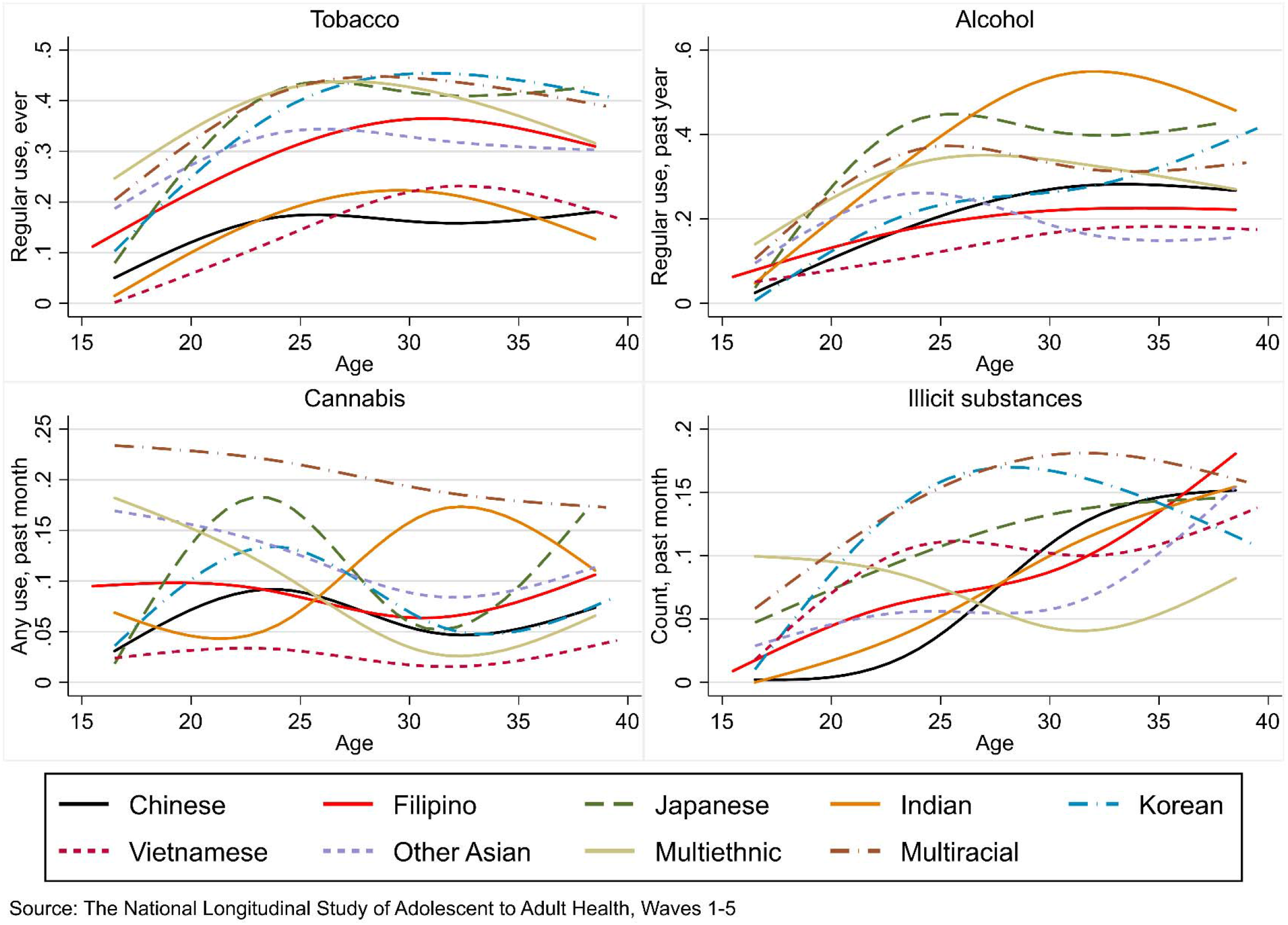
Smoothed substance use means by age and Asian ethnicity for tobacco, alcohol, cannabis, and illicit substances (median splines with 4 knots). Variable Y axes used to maximize acuity. (MI=30)

Figure 4 visualizes smoothed substance use means by age and generation-sex combination for tobacco, alcohol, cannabis, and illicit substances, using median splines as above. The expected sex and generation patterns are readily discernable, with males from generation 3+ exhibiting the highest level of use across all substances, and females from generation 1 exhibiting the lowest levels across substances. Interestingly, for illicit substances there is a pattern of convergence among nativity generations at the end of the age range, around age 35, while the gender disparity remains throughout.

**Figure 4:**
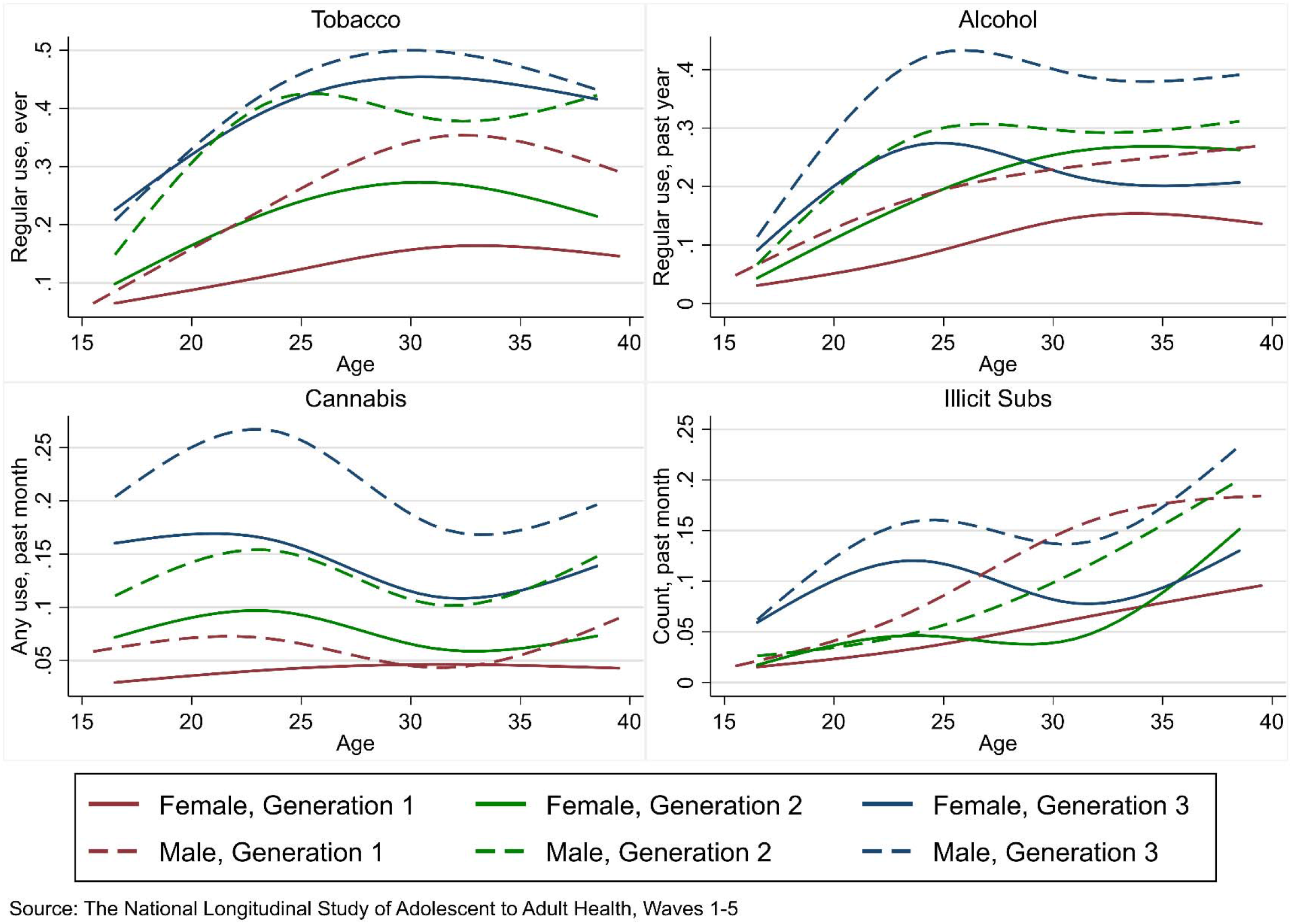
Smoothed substance use means by age, sex, and nativity generation for tobacco, alcohol, cannabis, and illicit substances (median splines with 4 knots). Variable Y axes used to maximize acuity. (MI=30)

#### Inferential analyses

Table 3, Models 1-4 examine ever daily tobacco use (at least 1 cigarette a day for a month) across Asian ethnicities, with Chinese ethnicity as the referent. Table 3, Model 1 shows that unconditional regular tobacco use varies substantially across Asian American ethnicities, with the highest probabilities of substance use among multiracial Asians (*β*=2.33; p<0.001) and multiethnic Asian (*β*=2.25; p<0.001), compared to the Chinese referent. Korean (*β*=2.15; p<0.001), Filipino (*β*=1.65; p<0.001), Japanese (*β*=1.65; p<0.01), and “Other” Asian ethnicities (*β*=2.09; p<0.001) also have higher unconditional probabilities of regular tobacco use, relative to Chinese Americans. In Table 3, Models 2 we control for sociodemographic variables: age, age squared, sex, and parental SES. Age and sex show independent effects on tobacco use in the expected direction; i.e., females are significantly less likely to be a daily tobacco user (*β*=-1.03; p<0.001) than males, and a curvilinear effect of age with tobacco use increasing at early ages (*β*=0.86; p<0.001) before tapering off at later ages (*β*=-0.01; p<0.001). All ethnic differences were substantively unchanged by sociodemographic covariate adjustment. Table 3, Model 3 considers contextual acculturation effects (i.e., co-ethnic peer network and neighborhood context), neither of which are significantly associated with tobacco use. In Table 3, Model 4, considers the influence of the generational acculturation variables—generational status (first (referent), second, and third+), and English language use in familial/private conversation. Results indicate AA from generations 2 and 3+ have significantly increased probability of tobacco use (p<0.001) relative to the first generation referent, as do AAs who use English in familial/private conversation (p<0.05). Adjusting for generational acculturation moderately attenuates the coefficients for high-risk ethnicities from Model 2, reducing the coefficients for multiethnic Asian by ~35%, multiracial Asian by ~40%, and “Other” Asian by ~20%); however, all of these coefficients remain significant. Conversely, controlling for acculturation factors attenuates the Japanese ethnicity coefficient by ~60%, reducing it to nonsignificance.

**Table 3.** Generalized linear mixed model coefficients (logistic link) predicting ever regular tobacco smoking, and regular alcohol use in the past year, by ethnicity, sociodemographics, and acculturation indicators across Add Health Waves 1-5 (MI=30)

| **<u>Ethnicity</u>** | Tobacco<br>Model 1 | Tobacco<br>Model 2 | Tobacco<br>Model 3 | Tobacco<br>Model 4 | Alcohol<br>Model 5 | Alcohol<br>Model 6 | Alcohol<br>Model 7 | Alcohol<br>Model 8 |
| --- | --- | --- | --- | --- | --- | --- | --- | --- |
| Chinese (ref.) | - | - | - | - | - | - | - | - |
| Filipino | 1.653***<br>(5.94) | 2.009***<br>(6.16) | 2.117***<br>(6.07) | 2.064***<br>(5.83) | -0.0780<br>(-0.47) | -0.162<br>(-0.86) | -0.0618<br>(-0.31) | -0.140<br>(-0.70) |
| Japanese | 1.647**<br>(2.72) | 2.015**<br>(2.74) | 2.127**<br>(2.88) | 0.816<br>(1.09) | 0.843*<br>(2.55) | 0.710<br>(1.91) | 0.741*<br>(1.99) | 0.415<br>(1.09) |
| Asian Indian | 0.658<br>(0.92) | 0.777<br>(0.91) | 0.741<br>(0.87) | 0.823<br>(0.96) | 0.951**<br>(2.66) | 1.008*<br>(2.53) | 0.917*<br>(2.29) | 0.846*<br>(2.15) |
| Korean | 2.147***<br>(4.86) | 2.847***<br>(5.25) | 2.846***<br>(5.24) | 2.741***<br>(5.04) | 0.496*<br>(2.02) | 0.640*<br>(2.33) | 0.678*<br>(2.47) | 0.680*<br>(2.46) |
| Vietnamese | 0.251<br>(0.48) | 0.346<br>(0.57) | 0.320<br>(0.53) | 0.570<br>(0.91) | -0.164<br>(-0.53) | 0.0219<br>(0.06) | 0.101<br>(0.29) | 0.184<br>(0.52) |
| Other Asian | 2.092***<br>(5.85) | 2.613***<br>(6.07) | 2.633***<br>(6.11) | 2.098***<br>(4.86) | -0.00961<br>(-0.05) | 0.0812<br>(0.35) | 0.0917<br>(0.40) | -0.0771<br>(-0.34) |
| Multiethnic Asian | 2.249***<br>(5.21) | 2.873***<br>(5.47) | 2.977***<br>(5.59) | 1.895***<br>(3.48) | 0.647**<br>(2.60) | 0.762**<br>(2.72) | 0.718*<br>(2.55) | 0.417<br>(1.46) |
| Multiracial Asian | 2.330***<br>(7.34) | 3.075***<br>(8.12) | 3.092***<br>(8.10) | 1.777***<br>(4.39) | 0.661***<br>(3.84) | 0.826***<br>(4.31) | 0.742***<br>(3.86) | 0.381<br>(1.81) |
| **<u>Sociodemographics</u>** |  |  |  |  |  |  |  |  |
| Age |  | 0.863***<br>(15.86) | 0.857***<br>(15.62) | 0.855***<br>(15.54) |  | 0.519***<br>(12.00) | 0.527***<br>(12.12) | 0.526***<br>(12.09) |
| Age squared |  | -0.014***<br>(-14.09) | -0.013***<br>(-13.85) | -0.013***<br>(-13.76) |  | -0.008***<br>(-10.08) | -0.008***<br>(-10.19) | -0.008***<br>(-10.16) |
| Sex (Female) |  | -1.032***<br>(-4.90) | -1.040***<br>(-4.94) | -0.987***<br>(-4.70) |  | -0.776***<br>(-6.94) | -0.775***<br>(-6.94) | -0.759***<br>(-6.84) |
| Parental Education |  | -0.0472<br>(-0.81) | -0.0537<br>(-0.92) | -0.0701<br>(-1.21) |  | 0.0648<br>(1.95) | 0.0634<br>(1.93) | 0.0530<br>(1.61) |
| Household Income |  | 0.151<br>(0.93) | 0.159<br>(0.97) | -0.0674<br>(-0.40) |  | 0.264**<br>(3.08) | 0.259**<br>(3.06) | 0.174*<br>(2.00) |
| **<u>Acculturation indicators</u>** |  |  |  |  |  |  |  |  |
| Co-ethnic tract proportion |  |  | -0.466<br>(-1.03) | -0.749<br>(-1.65) |  |  | 0.602*<br>(2.10) | 0.443<br>(1.53) |
| Co-ethnic friend network |  |  | -0.112<br>(-0.30) | 0.299<br>(0.80) |  |  | -0.609**<br>(-2.99) | -0.475*<br>(-2.30) |
| 1st gen (ref.) |  |  |  | - |  |  |  | - |
| 2nd gen |  |  |  | 1.088***<br>(3.52) |  |  |  | 0.339*<br>(2.06) |
| 3rd gen + |  |  |  | 2.211***<br>(6.95) |  |  |  | 0.447**<br>(2.60) |
| English use |  |  |  | 0.503*<br>(2.24) |  |  |  | 0.405*<br>(2.57) |
| Intercept | -3.851***<br>(-14.27) | -17.14***<br>(-17.85) | -16.90***<br>(-16.85) | -17.02***<br>(-16.90) | -2.091***<br>(-15.09) | -11.00***<br>(-16.03) | -10.87***<br>(-15.56) | -10.90***<br>(-15.58) |
| Random intercept variance | 8.994***<br>(10.27) | 15.81***<br>(8.95) | 15.91***<br>(8.94) | 14.46***<br>(9.02) | 1.492***<br>(8.78) | 1.918***<br>(8.82) | 1.887***<br>(8.71) | 1.789***<br>(8.63) |
| N | 1584 | 1584 | 1584 | 1584 | 1584 | 1584 | 1584 | 1584 |
| Obs | 7920 | 7920 | 7920 | 7920 | 7920 | 7920 | 7920 | 7920 |

Table 3, Models 5-8 examine current regular alcohol consumption (≥1-2 drinks weekly in the past year) across Asian ethnicities. Results indicate that Japanese (*β*=0.84; p<0.05), Asian Indian (*β*=0.95; p<0.01), and Korean (*β*=0.50; p<0.01) Americans, as well as multiethnic (*β*=0.65; p<0.01) and multiracial AAs (S=0.66; p<0.001), have higher probabilities of alcohol use than the Chinese referent. These coefficients remain significant with covariate adjustment for sociodemographics and contextual acculturation factors (i.e., Table 3, Models 7). Age and sex show significant independent effects on regular alcohol use in the expected direction. Childhood household income shows positive association to regular alcohol use (S=0.26; p<0.01). Inclusion of contextual acculturation variables in Table 3, Model 7 show a marginally significant positive effect for co-ethnic neighborhood proportion (*β*=0.60; p<0.05), and a significant negative effect for co-ethnic friend network proportion (*β*=-0.61; p<0.01). Table 3, Model 8 shows the expected positive effects of generational acculturation variables (English use and second generation: p<0.05; third+ generation: p<0.01). The inclusion of the acculturation variables in Model 8 attenuates, to nonsignificance, the coefficients for multiethnic (~40%) and multiracial (~50%) AAs.

In Table 4, Models 1-4, we examine cannabis use in the past month across Asian ethnicities. Table 4, Models 1 shows considerable variability in cannabis use across Asian ethnicities, with increased levels of cannabis use among multiracial *(β* =1.80; p<0.001), multiethnic (*β*=0.94; p<0.01), and “Other” (*β*=1.11; p<0.001) AAs, compared to Chinese referent. In Table 4, Model 2, we adjust for sociodemographic variables. Parental household income has a small positive, independent effect on cannabis use (*β*=0.24; p<0.05), females are less likely to use cannabis than males (*β*=-0.64; p<0.001), and age exhibits the expected curvilinear pattern (age: *β*=0.11, p<0.05; age squared: *β*=-0.002, p<0.01). We examine the effects of contextual acculturation variables in Table 4, Model 3, which show a negative effect for co-ethic peer proportion (*β*=-0.68; p<0.01). Finally, Table 4, Model 4 examines generational acculturation variables, which show significant, independent effects in the expected direction (second generation: p<0.05; third+ generation: p<0.001, English use: p<0.01), with greater levels of acculturation associated with higher probability of cannabis use. Inclusion of generational acculturation variables attenuates the multiethnic Asian effect by ~65%, and reduces it to nonsignificance.

**Table 4.** Generalized linear mixed model coefficients predicting cannabis use (logistic link) and illicit substance use (negative binomial link) in the past month, by ethnicity, sociodemographics, and acculturation indicators across Add Health Waves 1-5 (MI=30)

| **<u>Ethnicity</u>** | Cannabis<br>Model 1 | Cannabis<br>Model 2 | Cannabis<br>Model 3 | Cannabis<br>Model 4 | Illicit<br>Model 5 | Illicit<br>Model 6 | Illicit<br>Model 7 | Illicit<br>Model 8 |
| --- | --- | --- | --- | --- | --- | --- | --- | --- |
| Chinese (ref.) |  |  |  |  |  |  |  |  |
| Filipino | 0.485*<br>(2.08) | 0.459*<br>(1.96) | 0.570*<br>(2.27) | 0.478<br>(1.89) | 0.281<br>(1.57) | 0.252<br>(1.39) | 0.303<br>(1.59) | 0.209<br>(1.05) |
| Japanese | 0.603<br>(1.14) | 0.324<br>(0.61) | 0.361<br>(0.68) | -0.286<br>(-0.53) | 0.571<br>(1.47) | 0.612<br>(1.57) | 0.622<br>(1.57) | 0.365<br>(0.90) |
| Asian Indian | 0.950<br>(1.75) | 0.884<br>(1.64) | 0.782<br>(1.45) | 0.752<br>(1.42) | 0.630<br>(1.59) | 0.645<br>(1.63) | 0.586<br>(1.48) | 0.518<br>(1.30) |
| Korean | 0.463<br>(1.28) | 0.499<br>(1.38) | 0.548<br>(1.52) | 0.453<br>(1.24) | 0.453<br>(1.56) | 0.528<br>(1.84) | 0.545<br>(1.90) | 0.481<br>(1.65) |
| Vietnamese | -0.559<br>(-1.09) | -0.340<br>(-0.66) | -0.254<br>(-0.49) | -0.126<br>(-0.25) | 0.334<br>(1.06) | 0.349<br>(1.10) | 0.397<br>(1.25) | 0.421<br>(1.32) |
| Other Asian | 1.106***<br>(3.78) | 1.205***<br>(4.14) | 1.222***<br>(4.20) | 0.923**<br>(3.20) | 0.279<br>(1.21) | 0.265<br>(1.15) | 0.274<br>(1.18) | 0.143<br>(0.61) |
| Multiethnic Asian | 0.938**<br>(2.71) | 0.934**<br>(2.72) | 0.881*<br>(2.52) | 0.291<br>(0.82) | 0.295<br>(1.07) | 0.334<br>(1.20) | 0.301<br>(1.06) | 0.0553<br>(0.19) |
| Multiracial Asian | 1.798***<br>(7.16) | 1.856***<br>(7.43) | 1.765***<br>(7.02) | 1.079***<br>(4.05) | 0.706***<br>(3.47) | 0.768***<br>(3.77) | 0.715***<br>(3.47) | 0.427<br>(1.90) |
| **<u>Sociodemographics</u>** |  |  |  |  |  |  |  |  |
| Age |  | 0.110*<br>(2.36) | 0.121*<br>(2.58) | 0.122*<br>(2.58) |  | 0.176***<br>(3.98) | 0.180***<br>(4.07) | 0.179***<br>(4.04) |
| Age squared |  | -0.002*<br>(-2.45) | -0.002**<br>(-2.64) | -0.002**<br>(-2.63) |  | -0.002*<br>(-2.54) | -0.002**<br>(-2.62) | -0.002*<br>(-2.58) |
| Sex (Female) |  | -0.638***<br>(-4.35) | -0.647***<br>(-4.41) | -0.630***<br>(-4.37) |  | -0.440***<br>(-3.74) | -0.440***<br>(-3.74) | -0.441***<br>(-3.77) |
| Parental Education |  | 0.0619<br>(1.57) | 0.0611<br>(1.54) | 0.0476<br>(1.21) |  | -0.00501<br>(-0.18) | -0.00477<br>(-0.17) | -0.0143<br>(-0.49) |
| Household Income |  | 0.237*<br>(1.99) | 0.236<br>(1.96) | 0.125<br>(1.04) |  | -0.0350<br>(-0.45) | -0.0363<br>(-0.47) | -0.0759<br>(-0.97) |
| **<u>Acculturation indicators</u>** |  |  |  |  |  |  |  |  |
| Co-ethnic tract proportion |  |  | 0.650<br>(1.81) | 0.380<br>(1.06) |  |  | 0.345<br>(1.15) | 0.225<br>(0.74) |
| Co-ethnic friend network |  |  | -0.678**<br>(-2.67) | -0.428<br>(-1.69) |  |  | -0.338<br>(-1.75) | -0.249<br>(-1.26) |
| 1st gen (ref.) |  |  |  |  |  |  |  |  |
| 2nd gen |  |  |  | 0.476*<br>(2.32) |  |  |  | -0.0207<br>(-0.13) |
| 3rd gen + |  |  |  | 0.988***<br>(4.72) |  |  |  | 0.203<br>(1.17) |
| English use |  |  |  | 0.497**<br>(2.84) |  |  |  | 0.336*<br>(2.03) |
| Intercept | -3.677***<br>(-16.66) | -5.926***<br>(-7.59) | -5.832***<br>(-7.35) | -5.993***<br>(-7.59) | -3.172***<br>(-18.47) | -6.064***<br>(-9.15) | -5.997***<br>(-8.93) | -5.976***<br>(-8.86) |
| Random intercept variance | 3.062***<br>(8.43) | 2.911***<br>(8.29) | 2.873***<br>(8.25) | 2.625***<br>(8.17) | 0.836***<br>(5.79) | 0.892***<br>(6.27) | 0.870***<br>(6.14) | 0.842***<br>(6.01) |
| N | 1584 | 1584 | 1584 | 1584 | 1584 | 1584 | 1584 | 1584 |
| Obs | 7920 | 7920 | 7920 | 7920 | 7920 | 7920 | 7920 | 7920 |

In Table 4, Models 5-8, we examine illicit substance use, using a count variable calculated from number of illicit substances used in the previous month. Although illicit substance use is relatively uncommon across Asian ethnicities, Table 4, Model 5 shows there is a highly significant increased probability of illicit substance use among multiracial Asians (*β*=0.71; p<0.001), compared to the Chinese referent. In Table 4, Model 6, parental education and childhood household income show no significant effect on illicit substance use, whereas sex has a large, independent effect in the expected direction, and age exhibits a strong positive linear effect (*β*=0.18; p<0.001), with a smaller quadratic taper (*β*=-0.002; p<0.05). In Table 4, Model 7 contextual acculturation variables show no significant associations to illicit substance use. Among the generational acculturation variables in Table 4, Model 8, only English use shows a significant (positive) effect on illicit substance use (*β*=0.34; p<0.05). Adjusting for generational acculturation variables attenuates the multiracial Asian coefficient by ~40%, reducing it to nonsignificance.

Based on the age and age squared estimates from the final models of the series, we estimated the average age of peak use for each of the four substance use outcomes to determine if the peaks occur significantly later than age 25, which the approximate age peak in contemporary U.S. cohorts (Chen and Jacobson 2012). The mean age of peak substance use in this sample was 32.02 (95% CI: 31.23 - 32.82) for tobacco; 33.07 (95% CI: 31.75 - 34.38) for alcohol; 26.18 (95% CI: 23.04 – 29.34) for cannabis, and 44.87 (95% CI: 26.22 – 62.91) for illicit substances. These results are largely consistent with the unconditional visualizations displayed in Figures 3 and 4. Cannabis is the only substance for which we fail to reject the null hypothesis that substance use peaks at age 25 or earlier in the contemporary AA population (α = 0.05). Indeed, these results indicate that tobacco and alcohol use actually peak significantly after age 30 in this contemporary AA cohort (p < 0.05). Thus, these analyses demonstrate that AA substance use peaks substantially later than the typical U.S. pattern for tobacco, alcohol, and illicit drugs. That said, we note the large CIs for illicit substances, as well as the out of range estimate for age of peak illicit substance use, suggesting that this estimate is quite approximate.

## Discussion

This research contributes to an improved understanding of the social epidemiology of substance use in four primary ways: 1) disaggregating analyses by AA ethnicity, including explicitly considering those identifying as multiethnic or multiracial; 2) separately analyzing a range of legal and illicit substances; 3) investigating the effects of a range of acculturation indicators on AA substance use patterns; and 4) examining longitudinal patterns of AA substance use from early adolescence to mature adulthood. In addition to its inferential aims, the study also provides a wealth of descriptive data visualizations mapping the intersectional patterns in which ethnicity, nativity generation, and gender shape substance use in the contemporary AA population. Additionally, the three guiding hypotheses all received substantial support. That is, we find strong supports for Hypothesis 1—AA ethnicities at high risk for substance use are characterized by high levels of acculturation, indicated by low proportion first generation (e.g., Japanese), and multiethnic and multiracial family structures. Relatedly, we find strong support for Hypothesis 2, as conditioning on acculturation indicators attenuated some, but not all, of the observed ethnic disparities, particularly for multiracial and multiethnic AAs. Finally, we found mixed support for Hypothesis 3, as cannabis use in this longitudinal AA cohort peaks in the mid-20s (~age 26), while AA substance use peaks considerably later (>age 30) for the other three substances considered (tobacco, alcohol, and illicit substances).

### Multiethnic and multiracial Asian Americans

Multiracial and, to a lesser extent, multiethnic AAs exhibited consistently elevated substance use rates across the range of substances studied. The finding of persistent elevated substance use among multiracial AAs, relative to other Asian ethnicities, were robust across all analyses performed. Our findings regarding high rates of substance use among multiracial AAs is novel, though there exist some findings on multiracial youth (not Asians) being at greater risks of alcohol consumption (Choi et al. 2006) and illicit substance use (Subica and Wu 2018). Underlying determinants of this high propensity of substance use among multiracial Asians is likely a high degree of acculturation. Although we have not tested the effects of psychological aspects, stress due to discordance between Asian and U.S. culture, as well as racial/ethnic discrimination (Brown 2018), likely contribute to the relatively high propensity for substance use observed for multiracial AAs.

Similarly, Asians who reported multiethnic Asian backgrounds exhibited a significantly higher probability of substance use, in 3 of the 4 substances studied than the Chinese referent. This finding, in part, reflects the effect of greater acculturation, as multiethnic AAs are more frequently third generation and English language users. Indeed, in the current study, 71% of multiethnic Asians identify as third generation (see Table 2), and 88% use English in familial/private conversation. It also seems reasonable to speculate that interethnic marriage may facilitate faster acculturation to U.S. culture, eroding the influence of each parent’s protective heritage culture in ways that go beyond English usage and personal and parental nativity. However, contrary to their general pattern of elevated substance use, multiethnic Asians exhibited relatively low levels of illicit substance use

### Acculturation and ethnicity

We found extensive support for Hypothesis 2. Conditioning on acculturation measures substantially attenuates the associations of ethnicity and substance use, with pronounced and consistent attenuation for multiracial and multiethnic Asians, as well as Japanese Americans. Indeed, the association of multiracial Asian identity to alcohol and illicit substance use were both rendered nonsignificant by adjusting for acculturation factors, as were the associations of multiethnic Asian identity to cannabis and alcohol, and Japanese ethnicity to tobacco and alcohol. However, analyses indicate that the association of substance use to ethnicity cannot be fully explained by acculturation. For instance, Korean ethnicity was robustly associated with increased tobacco and alcohol use conditional on all acculturation and sociodemographic controls. Additionally, co-ethnic peer networks demonstrated some independent, protective effects, particularly for alcohol use, which contributes to the growing literature describing the salubrious effects of Asian co-ethnic peer context (Ryabov 2015, Saraiya et al. 2019).

### Elevated tobacco use across a diverse range of Asian American ethnicities

Findings on tobacco smoking are statistically significant across several ethnic groups, suggesting significant variation in smoking prevalence among Asian groups. As illustrated in Figure 1, multiethnic Asians and multiracial Asians are at the highest risk among AA groups in terms of smoking. Even adjusting for relatively high acculturation, multiethnic and multiracial Asians still show a significantly higher probability of smoking than subjects of Chinese ethnicity. Additionally, rates of smoking are notably high among those of Korean ethnicity, who, despite having a high proportion of first generation immigrants, still exhibit significantly higher smoking rates than the Chinese referent. Elevated smoking rates are also seen for Japanese, Filipino, and “Other” Asian ethnicities relative to the Chinese referent. The high prevalence in tobacco smoking across a range of East and Southeast Asian nations of origin likely explain the observed elevated rates for these ethnicities (WHO 2015). These findings regarding tobacco use support previous findings demonstrating that the relationship between Asian ethnicity and substance use, in part, reflects cultural differences in nations of origin (Cook et al. 2015).

### Variation in age patterns of Asian American substance use

We found partial support for Hypothesis 3, which predicted that AA substance use would peak later than the U.S. average (i.e., approximately age 25, (Chen and Jacobson 2012)). Three of the four substances demonstrated peaks substantially later than age 25 – age 32 for tobacco, 33 for alcohol, and 45 for illicit substance use. Only cannabis exhibited an estimated age of peak use with a 95% CI overlapping age 25 (26.18; 95% CI: 23.04 – 29.34). Disturbingly, illicit substance use increases almost linearly across the entire period (ages ~15-40), exhibiting steady growth across the entire age range, 15-40, as visualized in Figure 3 and 4. All four substances exhibited a curvilinear functional form for age, in which substance use increases at earlier ages, and then declines (or decelerates, in the case of illicit substances at later ages. Thus, relative to the pattern observed in the U.S. as a whole (Chen and Jacobson 2012), AA substance use appears to peak at relatively late ages. Future research in independent samples will be needed to replicate this findings, as well as to disentangle to influences of age/period/cohort on the observed patterns.

### Limitations and future directions

Notwithstanding the current study’s contributions, it was characterized by various limitations. For instance, substance use measures in the study are based on self-reports; a notable, common, limitation in substance use survey research, as respondents tend to underreport substance use, due to social desirability bias (Ryabov 2015). Another limitation prevalent in longitudinal studies spanning multiple developmental stages is measurement inconsistency across waves, which was present here in the “illicit substance use” measure. Another limitations is the heterogeneous time periods of the four substance use outcomes (e.g., ever regular tobacco smoker, regular alcohol user in past year, cannabis use in the past month, illicit substance use in the past month), which was necessitated by the data available in Add Health. Lastly, small cell size in Asian Indian and Japanese respondents yielded large standard errors, limiting our ability to make statistical inferences for these groups. Future research into the social epidemiology of substance misuse would benefit from collecting larger longitudinal AA cohorts, and from considering intersectional variation across sex, ethnicity, sexual orientation, and acculturation statuses (e.g., co-ethnic marriage/homogamy). Additionally, future studies would benefit from modeling the role of polysubstance use among AAs.

## Conclusion

This study indicates significant substance use disparities across Asian ethnicities, with the lowest probability of substance use among Chinese and Vietnamese, and the highest among multiethnic and multiracial Asians. Moreover, acculturation indicators (e.g., English language use, nativity, and parental nativity) were strongly associated with increased substance use, and attenuate a degree of the observed ethnic disparities, particularly for multiracial and multiethnic Asians. This research demonstrates how the model minority stereotype may masks significant ethnic disparities within the Asian American population, and argues that a considerable proportion of the observed ethnic disparities in substance use stems from diverse family ties speeding transition toward U.S. cultural norms, and away from the relatively salubrious norms of Asian nations of origin.

## Data Availability

This research uses data from Add Health, a program project directed by Kathleen Mullan Harris and designed by J. Richard Udry, Peter S. Bearman, and Kathleen Mullan Harris at the University of North Carolina at Chapel Hill, and funded by grant P01-HD31921 from the Eunice Kennedy Shriver National Institute of Child Health and Human Development, with cooperative funding from 23 other federal agencies and foundations. Special acknowledgment is due to Ronald R. Rindfuss and Barbara Entwisle for assistance in the original design. Information on how to obtain the Add Health data files is available on the Add Health website (http://www.cpc.unc.edu/addhealth). No direct support was received from grant P01-HD31921 for this analysis.

## Compliance with Ethical Standards

All authors (i.e., Zobayer Ahmmad and Daniel E. Adkins) declare they have no conflicts of interest.

## Acknowledgments

We thank the Consortium of Families and Health Research (C-FAHR) at the University of Utah for providing access to the data. We are grateful to Bethany G Everett for editorial input. This research uses data from Add Health, a program project directed by Kathleen Mullan Harris and designed by J. Richard Udry, Peter S. Bearman, and Kathleen Mullan Harris at the University of North Carolina at Chapel Hill, and funded by grant P01-HD31921 from the Eunice Kennedy Shriver National Institute of Child Health and Human Development, with cooperative funding from 23 other federal agencies and foundations. Special acknowledgment is due to Ronald R. Rindfuss and Barbara Entwisle for assistance in the original design. Information on how to obtain the Add Health data files is available on the Add Health website (http://www.cpc.unc.edu/addhealth). No direct support was received from grant P01-HD31921 for this analysis.

---

**Appendix 1:** Missing data in the sample

| Variable | Missing | Total | % Missing |
| --- | --- | --- | --- |
| W1 Tobacco | 2 | 1,584 | 0.13 |
| W1 Alcohol | 3 | 1,584 | 0.19 |
| W1 Cannabis | 26 | 1,584 | 1.64 |
| W1 Illicit subs. | 20 | 1,584 | 1.26 |
| W2 Tobacco | 495 | 1,584 | 31.25 |
| W2 Alcohol | 495 | 1,584 | 31.25 |
| W2 Cannabis | 512 | 1,584 | 32.32 |
| W2 Illicit subs. | 497 | 1,584 | 31.38 |
| W3 Tobacco | 395 | 1,584 | 24.94 |
| W3 Alcohol | 400 | 1,584 | 25.25 |
| W3 Cannabis | 393 | 1,584 | 24.81 |
| W3 Illicit subs. | 392 | 1,584 | 24.75 |
| W4 Tobacco | 524 | 1,584 | 33.08 |
| W4 Alcohol | 524 | 1,584 | 33.08 |
| W4 Cannabis | 524 | 1,584 | 33.08 |
| W4 Illicit subs. | 524 | 1,584 | 33.08 |
| W5 Tobacco | 729 | 1,584 | 46.02 |
| W5 Alcohol | 727 | 1,584 | 45.9 |
| W5 Cannabis | 729 | 1,584 | 46.02 |
| W5 Illicit subs. | 730 | 1,584 | 46.09 |
| Chinese | 0 | 1,584 | 0 |
| Filipino | 0 | 1,584 | 0 |
| Japanese | 0 | 1,584 | 0 |
| Asian Indian | 0 | 1,584 | 0 |
| Korean | 0 | 1,584 | 0 |
| Vietnamese | 0 | 1,584 | 0 |
| Other Asian | 0 | 1,584 | 0 |
| Multiethnic Asian | 0 | 1,584 | 0 |
| Multiracial Asian | 0 | 1,584 | 0 |
| W1 Age | 0 | 1,584 | 0 |
| Sex (Female) | 0 | 1,584 | 0 |
| Household Income (Logged) | 673 | 1,584 | 42.49 |
| Parental education (mean) | 88 | 1,584 | 5.56 |
| Generation 1 | 221 | 1,584 | 13.95 |
| Generation 2 | 178 | 1,584 | 11.24 |
| Generation 3+ | 220 | 1,584 | 13.89 |
| W1 English use | 0 | 1,584 | 0 |
| W2 English use | 496 | 1,584 | 31.31 |
| W3 English use | 389 | 1,584 | 24.56 |
| W1 Co-ethnic tract proportion | 9 | 1,584 | 0.57 |
| W2 Co-ethnic tract proportion | 505 | 1,584 | 31.88 |
| W3 Co-ethnic tract proportion | 397 | 1,584 | 25.06 |
| W4 Co-ethnic tract proportion | 524 | 1,584 | 33.08 |
| Co-ethnic friend network | 585 | 1,584 | 36.93 |

Note: No data were collected for language use variables in Waves 4 & 5 and co-ethnic tract proportion in Wave 5.

## References

Abraído-Lanza, Ana F., Maria T. Chao and Karen R. Florez. 2005. “Do Healthy Behaviors Decline with Greater Acculturation?: Implications for the Latino Mortality Paradox.” Social science & medicine 61(6):1243–55.

Author(s) - blinded for peer review

Becerra, Monideepa B., Patti Herring, Helen Hopp Marshak and Jim E. Banta. 2013. “Association between Acculturation and Binge Drinking among Asian-Americans: Results from the California Health Interview Survey.” Journal of Addiction 2013. doi: 10.1155/2013/248196.

Bersamira, Clifford S., Yu-An Lin, Keunhye Park and Jeanne C. Marsh. 2017. “Drug Use among Asian Americans: Differentiating Use by Acculturation Status and Gender.” Journal of Substance Abuse Treatment 79:76–81. doi: 10.1016/j.jsat.2017.06.002.

Bollen, Kenneth A., Paul P. Biemer, Alan F. Karr, Stephen Tueller and Marcus E. Berzofsky. 2016. “Are Survey Weights Needed? A Review of Diagnostic Tests in Regression Analysis.” Annual Review of Statistics and Its Application 3(1):375–92. doi: 10.1146/annurev-statistics-011516-012958.

Brown, Tyson H. 2018. “Racial Stratification, Immigration, and Health Inequality: A Life Course-Intersectional Approach.” Social Forces 96(4):1507–40. doi: 10.1093/sf/soy013 %J Social Forces.

Case, Anne and Angus Deaton. 2015. “Rising Morbidity and Mortality in Midlife among White Non-Hispanic Americans in the 21st Century.” Proceedings of the National Academy of Sciences 112(49):15078. doi: 10.1073/pnas.1518393112.

Chen, P. and K. C. Jacobson. 2012. “Developmental Trajectories of Substance Use from Early Adolescence to Young Adulthood: Gender and Racial/Ethnic Differences.” Journal of Adolescent Health 50(2):154–63. doi: 10.1016/j.jadohealth.2011.05.013.

Choi, Yoonsun, Tracy W. Harachi, Mary Rogers Gillmore and Richard F. Catalano. 2006. “Are Multiracial Adolescents at Greater Risk? Comparisons of Rates, Patterns, and Correlates of Substance Use and Violence between Monoracial and Multiracial Adolescents.” American Journal of Orthopsychiatry 76(1):86–97. doi: 10.1037/0002-9432.76.1.86.

Cochran, Susan D., Vickie M. Mays, Margarita Alegria, Alexander N. Ortega and David Takeuchi. 2007. “Mental Health and Substance Use Disorders among Latino and Asian American Lesbian, Gay, and Bisexual Adults.” Journal of Consulting and Clinical Psychology 75(5):785–94. doi: 10.1037/0022-006X.75.5.785.

Connor, J. P., M. J. Gullo, A. White and A. B. Kelly. 2014. “Polysubstance Use: Diagnostic Challenges, Patterns of Use and Health.” Current Opinion in Psychiatry 27(4):269–75. doi: 10.1097/yco.0000000000000069.

Cook, Won Kim, Nina Mulia and Katherine Karriker-Jaffe. 2012. “Ethnic Drinking Cultures and Alcohol Use among Asian American Adults: Findings from a National Survey.” Alcohol and Alcoholism 47(3):340–48. doi: 10.1093/alcalc/ags017.

Cook, Won Kim, Jason Bond, Katherine J. Karriker-Jaffe and Sarah Zemore. 2013. “Who’s at Risk? Ethnic Drinking Cultures, Foreign Nativity, and Problem Drinking among Asian American Young Adults.” Journal of Studies on Alcohol and Drugs 74(4):532–41. doi: 10.15288/jsad.2013.74.532.

Cook, Won Kim, Katherine J. Karriker-Jaffe, Jason Bond and Camillia Lui. 2015. “Asian American Problem Drinking Trajectories During the Transition to Adulthood: Ethnic Drinking Cultures and Neighborhood Contexts.” American journal of public health 105(5):1020. doi: 10.2105/AJPH.2014.302196.

Cook, Won Kim, Winston Tseng, Christina Tam, Iyanrick John and Camillia Lui. 2017. “Ethnic-Group Socioeconomic Status as an Indicator of Community-Level Disadvantage: A Study of Overweight/Obesity in Asian American Adolescents.” Soc Sci Med 184:15–22. doi: 10.1016/j.socscimed.2017.04.027.

Entzel, P. and J. R. Udry. 2009. “The National Longitudinal Study of Adolescent to Adult Health: Research Design [Www Document].”

Fothergill, Kate, Margaret E. Ensminger, Elaine E. Doherty, Hee-Soon Juon and Kerry M. Green. 2016. “Pathways from Early Childhood Adversity to Later Adult Drug Use and Psychological Distress: A Prospective Study of a Cohort of African Americans.” Journal of Health and Social Behavior 57(2):223–39. doi: 10.1177/0022146516646808.

Gelman, A. and J. Hill. 2006. Data Analysis Using Regression and Multilevel/Hierarchical Models: Cambridge University Press.

Geronimus, A. T., J. Bound, T. A. Waidmann, J. M. Rodriguez and B. Timpe. 2019. “Weathering, Drugs, and Whack-a-Mole: Fundamental and Proximate Causes of Widening Educational Inequity in Us Life Expectancy by Sex and Race, 1990–2015.” Journal of Health and Social Behavior 60(2):222’39. doi: 10.1177/0022146519849932.

Hahm, Hyeouk, Judith Gonyea, Christine Chiao and Luca Koritsanszky. 2014. “Fractured Identity: A Framework for Understanding Young Asian American Women’s Self-Harm and Suicidal Behaviors.” Race and Social Problems 6(1):56–68. doi: 10.1007/s12552-014-9115-4.

Hahm, Hyeouk Chris, Frank Y. Wong, Zhihuan Jennifer Huang, Al Ozonoff and Jieha Lee. 2008. “Substance Use among Asian Americans and Pacific Islanders Sexual Minority Adolescents: Findings from the National Longitudinal Study of Adolescent Health.” Journal of Adolescent Health 42(3):275–83. doi: 10.1016/j.jadohealth.2007.08.021.

Hong, Jun Sung, Hui Huang, Bushra Sabri and Johnny S. Kim. 2011. “Substance Abuse among Asian American Youth: An Ecological Review of the Literature.” Children and Youth Services Review 33(5):669–77. doi: 10.1016/j.childyouth.2010.11.015.

Iwamoto, Derek, Stephanie Takamatsu and Jeanett Castellanos. 2012. “Binge Drinking and Alcohol-Related Problems among U.S.-Born Asian Americans.” Cultural Diversity and Ethnic Minority Psychology 18(3):219–27. doi: 10.1037/a0028422.

Iwamoto, Derek Kenji, Aylin Kaya, Margaux Grivel and Lauren Clinton. 2016. “Under-Researched Demographics: Heavy Episodic Drinking and Alcohol-Related Problems among Asian Americans.” Alcohol research : current reviews 38(1):17.

Kline, David, Rebecca Andridge and Eloise Kaizar. 2017. “Comparing Multiple Imputation Methods for Systematically Missing Subject-Level Data.” Research Synthesis Methods 8(2):136–48. doi: 10.1002/jrsm.1192.

Kochhar, Rakesh and Anthony Cilluffo. 2018. “Income Inequality in the Us Is Rising Most Rapidly among Asians.”.” Pew Research Center report, Social and Demographic Trends, July 12.

Le, Thao, Deborah Goebert and Judy Wallen. 2009. “Acculturation Factors and Substance Use among Asian American Youth.” The Journal of Primary Prevention 30(3):453–73. doi: 10.1007/s10935-009-0184-x.

Le, Thao N., and Tomoko Kato. 2006. “The Role of Peer, Parent, and Culture in Risky Sexual Behavior for Cambodian and Lao/Mien Adolescents.” Journal of Adolescent Health 38(3):288–96. doi: 10.1016/j.jadohealth.2004.12.005.

Lee, Jennifer and Frank D. Bean. 2004. “America’s Changing Color Lines: Immigration, Race/Ethnicity, and Multiracial Identification.” Annu. Rev. Sociol. 30: 221–42.

Lee, Jennifer. 2015. The Asian American Achievement Paradox, Edited by M. Zhou: New York: Russell Sage Foundation.

Lee, Katherine J., and John B. Carlin. 2010. “Multiple Imputation for Missing Data: Fully Conditional Specification Versus Multivariate Normal Imputation.” American Journal of Epidemiology 171(5):624–32. doi: 10.1093/aje/kwp425 %J American Journal of Epidemiology.

Li, Kelin and Ming Wen. 2015. “Substance Use, Age at Migration, and Length of Residence among Adult Immigrants in the United States.” Journal of Immigrant and Minority Health 17(1):156–64. doi: 10.1007/s10903-013-9887-4.

Liu, William Ming and Derek Kenji Iwamoto. 2007. “Conformity to Masculine Norms, Asian Values, Coping Strategies, Peer Group Influences and Substance Use among Asian American Men.” Psychology of Men & Masculinity 8(1):25–39. doi: 10.1037/1524-9220.8.1.25.

López, Gustavo, Neil G. Ruiz and Eileen Patten. 2017. “Key Facts About Asian Americans, a Diverse and Growing Population.” Pew Research Center. Accessed February 9: 2018.

Lui, P. Priscilla and Byron L. Zamboanga. 2018. “Acculturation and Alcohol Use among Asian Americans: A Meta-Analytic Review.” Psychology of Addictive Behaviors 32(2):173–86. doi: 10.1037/adb0000340.

Martell, Brandi N., Bridgette E. Garrett and Ralph S. Caraballo. 2016. “Disparities in Adult Cigarette Smoking--United States, 2002–2005 and 2010-2013.” Morbidity and Mortality Weekly Report 65(30):753. doi: 10.15585/mmwr.mm6530a1.

McHugh, R. K., V. R. Votaw, D. E. Sugarman and S. F. Greenfield. 2018. “Sex and Gender Differences in Substance Use Disorders.” Clinical Psychology Review 66:12–23. doi: 10.1016/j.cpr.2017.10.012.

Merline, Alicia C., Patrick M. O’Malley, John E. Schulenberg, Jerald G. Bachman and Lloyd D. Johnston. 2004. “Substance Use among Adults 35 Years of Age: Prevalence, Adulthood Predictors, and Impact of Adolescent Substance Use.” The American Journal of Public Health 94(1):96. doi: 10.2105/AJPH.94.1.96.

Nelson, Sarah C., Nazneen F. Bahrassa, Moin Syed and Richard M. Lee. 2015. “Transitions in Young Adulthood: Exploring Trajectories of Parent–Child Conflict During College.” Journal of Counseling Psychology 62(3):545–51. doi: 10.1037/cou0000078.

Oehlert, Gary W. 1992. “A Note on the Delta Method.” The American Statistician 46(1):27–29. doi: 10.1080/00031305.1992.10475842.

Park, So-Youn, Jeane Anastas, Tazuko Shibusawa and Duy Nguyen. 2014. “The Impact of Acculturation and Acculturative Stress on Alcohol Use across Asian Immigrant Subgroups.” Substance Use & Misuse, 2014, Vol.49(8), p.922–931 49(8):922–31. doi: 10.3109/10826084.2013.855232.

Qian, Zhenchao and Daniel T. Lichter. 2011. “Changing Patterns of Interracial Marriage in a Multiracial Society.” Journal of Marriage and Family 73(5):1065–84. doi: 10.1111/j.1741-3737.2011.00866.x.

Royston, Patrick and Willi Sauerbrei. 2007. “Multivariable Modeling with Cubic Regression Splines: A Principled Approach.” The Stata Journal 7(1):45–70. doi: 10.1177/1536867X0700700103.

Ruhm, C. J. 2017. “Geographic Variation in Opioid and Heroin Involved Drug Poisoning Mortality Rates.” American Journal of Preventive Medicine 53(6):745–53. doi: 10.1016/j.amepre.2017.06.009.

Ruhm, C. J. 2018. “Drug Mortality and Lost Life Years among Us Midlife Adults, 1999–2015.” American Journal of Preventive Medicine 55(1):11–18. doi: 10.1016/j.amepre.2018.03.014.

Ryabov, Igor. 2015. “Relation of Peer Effects and School Climate to Substance Use among Asian American Adolescents.” Journal of Adolescence 42(C):115–27. doi: 10.1016/j.adolescence.2015.04.007.

Saraiya, T., K. Z. Smith, A. N. C. Campbell and D. Hien. 2019. “Posttraumatic Stress Symptoms, Shame, and Substance Use among Asian Americans.” Journal of Substance Abuse Treatment 96:1–11. doi: 10.1016/j.jsat.2018.10.002.

Schwartz, S. J., J. B. Unger, B. L. Zamboanga and J. Szapocznik. 2010. “Rethinking the Concept of Acculturation Implications for Theory and Research.” American Psychologist 65(4):237–51. doi: 10.1037/a0019330.

Shanahan, L., S. N. Hill, L. M. Gaydosh, A. Steinhoff, E. J. Costello, K. A. Dodge, K. M. Harris and W. E. Copeland. 2019. “Does Despair Really Kill? A Roadmap for an Evidence-Based Answer.” American Journal of Public Health 109(6):854–58. doi: 10.2105/ajph.2019.305016.

Subica, Andrew M., and Li-Tzy Wu. 2018. “Substance Use and Suicide in Pacific Islander, American Indian, and Multiracial Youth.” American Journal of Preventive Medicine 54(6):795–805. doi: 10.1016/j.amepre.2018.02.003.

Telzer, Eva, Nancy Gonzales and Andrew Fuligni. 2014. “Family Obligation Values and Family Assistance Behaviors: Protective and Risk Factors for Mexican-American Adolescents’ Substance Use.” Journal of Youth and Adolescence 43(2):270–83. doi: 10.1007/s10964-013-9941-5.

Thai, Nghi D., Christian M. Connell and Jacob Kraemer Tebes. 2010. “Substance Use among Asian American Adolescents: Influence of Race, Ethnicity, and Acculturation in the Context of Key Risk and Protective Factors.” Asian American Journal of Psychology 1(4):261–74. doi: 10.1037/a0021703.

Tosh, Aneesh K., and Patricia S. Simmons. 2007. “Sexual Activity and Other Risk-Taking Behaviors among Asian-American Adolescents.” Journal of Pediatric and Adolescent Gynecology 20(1):29–34. doi: 10.1016/j.jpag.2006.10.010.

von Hippel, Paul T. 2018. “How Many Imputations Do You Need? A Two-Stage Calculation Using a Quadratic Rule.” Sociological Methods & Research: In press. https://doi.org/10.1177/0049124117747303. doi: 10.1177/0049124117747303.

White, Ian R, Patrick Royston and Angela M Wood. 2011. “Multiple Imputation Using Chained Equations: Issues and Guidance for Practice.” Statistics in Medicine 30(4):377–99.

WHO. 2019. “Report on the Global Tobacco Epidemic, 2019.” Vol. CC BY-NC-SA 3.0 IGO. Geneva: World Health Organization.

WHO, World Health Organization. 2015. Who Global Report on Trends in Prevalence of Tobacco Smoking 2015: World Health Organization.

Yoo, Grace J., Mai-Nhung Le and Alan Y. Oda. 2013. Handbook of Asian American Health: New York: Springer.

Yoo, Hyung, Gilbert Gee, Craig Lowthrop and Joanne Robertson. 2010. “Self-Reported Racial Discrimination and Substance Use among Asian Americans in Arizona.” Journal of Immigrant and Minority Health 12(5):683–90. doi: 10.1007/s10903-009-9306-z.

